# Long chain omega-3 fatty acid intake in pregnancy and risk of type 1 diabetes in the offspring: Two large Scandinavian pregnancy cohorts – MoBa and DNBC

**DOI:** 10.1101/2023.11.23.23297665

**Authors:** Nicolai A. Lund-Blix, Anne A. Bjerregaard, German Tapia, Ketil Størdal, Anne Lise Brantsæter, Marin Strøm, Thorhallur I. Halldorsson, Charlotta Granstrøm, Jannet Svensson, Geir Joner, Torild Skrivarhaug, Pål R. Njølstad, Sjurdur F. Olsen, Lars C. Stene

## Abstract

**Background:** Long-chain marine omega-3 fatty acids (eicosapentaenoic acid; EPA and docosahexaenoic acid; DHA) have anti-inflammatory effects. Dietary intake of EPA and DHA in pregnancy was associated with lower offspring risk of asthma in a randomized trial, and lower risk of type 1 diabetes in the offspring in retrospective observational studies.

**Aim:** We aimed to investigate whether higher intakes of EPA and DHA during pregnancy is associated with a lower type 1 diabetes risk in children in two of the world’s largest birth cohorts.

**Methods:** The Danish National Birth Cohort (DNBC) and the Norwegian Mother, Father and Child Cohort Study (MoBa) together include 153,843 mother-child pairs with prospectively collected data on EPA and DHA intake during pregnancy using validated food frequency questionnaires. Type 1 diabetes diagnosis in children (n=634) was ascertained from national diabetes registries.

**Results:** There was no association between the sum of EPA and DHA intake during pregnancy and offspring risk of type 1 diabetes (hazard ratio per g/d of intake: 1.00 in both MoBa and DNBC, pooled 95% CI: 0.88-1.14). Adjustment for potential cofounders and robustness analyses gave very similar results.

**Conclusion:** The hypothesis that a higher maternal omega-3 fatty acid intake during pregnancy reduce the risk of offspring type 1 diabetes was not supported.

## Introduction

Long-chain marine omega-3 fatty acids (eicosapentaenoic acid [EPA] and docosahexaenoic acid [DHA]) have anti-inflammatory effects (1, 2) and may influence the development of type 1 diabetes (3). Studies in experimental animals (4) and retrospective case-control studies (5) have reported associations between early life intake of omega-3 fatty acids and lower risk of type 1 diabetes. In the spontaneously diabetic NOD-mouse model, dietary intervention with EPA + DHA reduced the incidence of type 1 diabetes and modulated inflammation and immunity including regulatory- and helper T-cells (4). Furthermore, high risk cohorts have reported associations between childhood intake of dietary fatty acids and risk of islet autoimmunity or type 1 diabetes, albeit only suggestive for EPA and DHA (3, 6, 7).

DPA, EPA and other fatty acids cross the placenta (8), and maternal dietary intake of these fatty acids during pregnancy may influence offspring physiology and health (9). Notably, a randomized trial of fish oil supplement (containing EPA and DHA) from the second trimester of pregnancy reduced the risk of asthma or wheezing in the children at five years of age (10), showing proof of concept that maternal DPA and DHA can influence the offspring risk of an immune-mediated disease. Few prospective studies have investigated maternal intake of marine omega-3 fatty acids in relation to risk of childhood onset type 1 diabetes (11).

We aimed to investigate whether higher intakes of the long chain marine n-3 fatty acids (EPA and DHA) in the maternal diet during pregnancy is associated with a lower risk of childhood onset type 1 diabetes in two of the largest birth cohorts in the world.

## Methods

The Danish National Birth Cohort (DNBC) recruited pregnant women from 1996-2002 and the Norwegian Mother, Father and Child Cohort Study (MoBa) from 1999–2008 (12, 13). Both cohorts are based on written consent and received ethical approval (Supplementary Methods). Dietary intake was assessed during pregnancy using validated harmonized food frequency questionnaires (Supplementary Methods), and the two cohorts included 153,843 mother-child pairs with data on maternal intake of EPA and DHA (Figure 1). We ascertained date of type 1 diabetes diagnosis for the children from the national childhood diabetes registries (excluding monogenic and type 2 diabetes) and used time from birth to diagnosis in adjusted Cox regression analyses. We used the sum of EPA and DHA intake from food and supplements during pregnancy as a continuous variable in the primary analyses, and we meta-analyzed the regression results from the two cohorts (details in Supplementary Methods).

**Figure 1.**
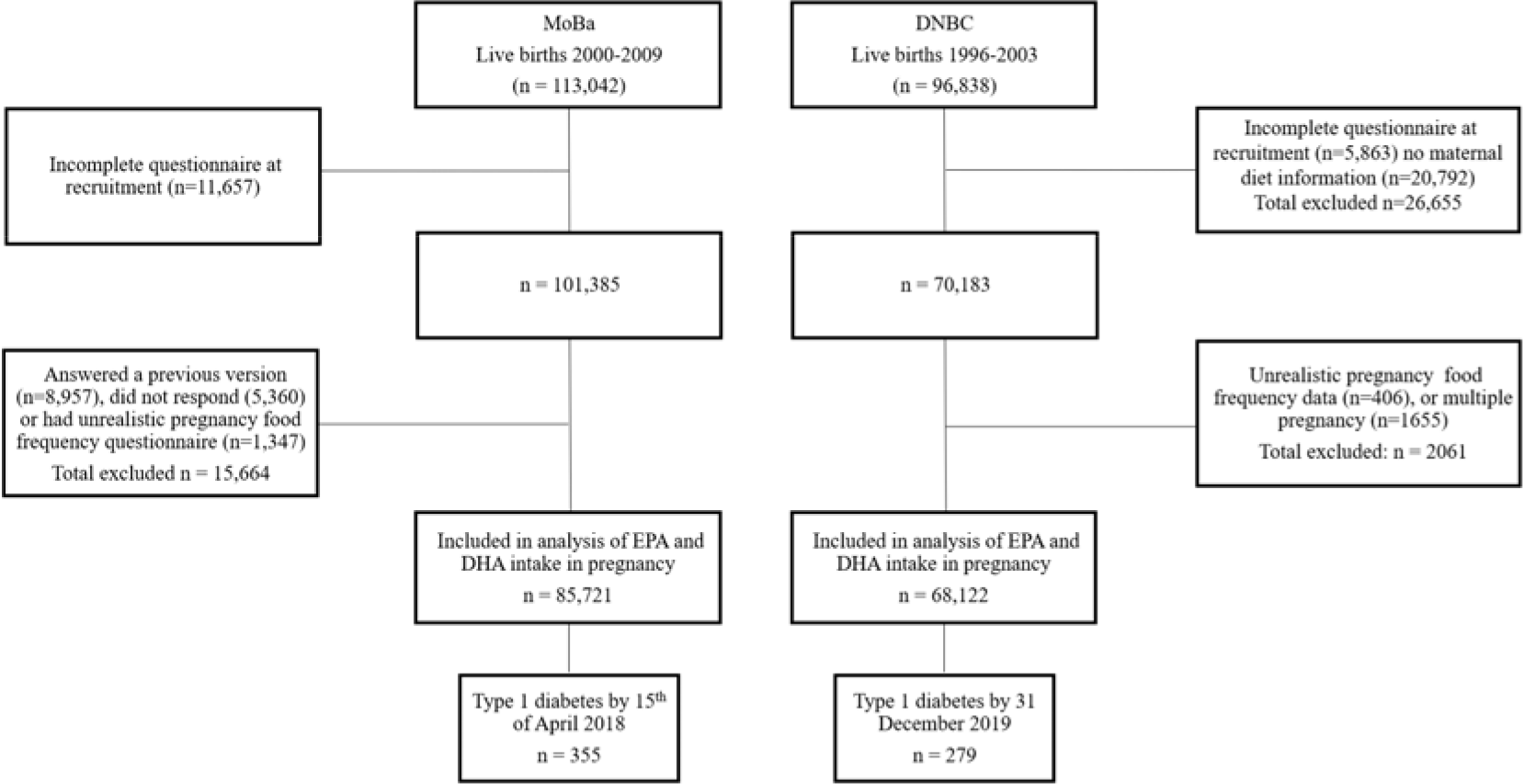
Flow chart for the generation of the analysis sample. DNBC: The Danish National Birth Cohort; MoBa (Norwegian acronym for): The Norwegian Mother, Father and Child Cohort Study. Total number of mother/child-pairs included in analysis: 153,843 (n=634 of whom developed type 1 diabetes).

## Results

Among 153,843 children, 634 (0.41%) developed type 1 diabetes during follow-up. The median total maternal intakes (and interquartile range) of sum EPA and DHA during pregnancy were 0.26 g/d (0.15 – 0.44) in DNBC and 0.56 g/d (0.32 – 1.07) in MoBa, with a higher intake of both fish and supplements in Norway compared to Denmark (Supplementary Table 1). Of the women, 46-48 % were nulliparous, 9-14% smoked during pregnancy and 68-77% had pre-pregnancy body mass index below 25 kg/m^2^ (Supplementary Table 2).

There was no association between the sum of EPA and DHA intake during pregnancy and the child’s risk of type 1 diabetes (hazard ratio 1.00 after adjustment for maternal parity, maternal pre-pregnancy BMI, maternal smoking during pregnancy, maternal type 1 diabetes, offspring sex, duration of breastfeeding and maternal education, Figure 2).

**Figure 2.**
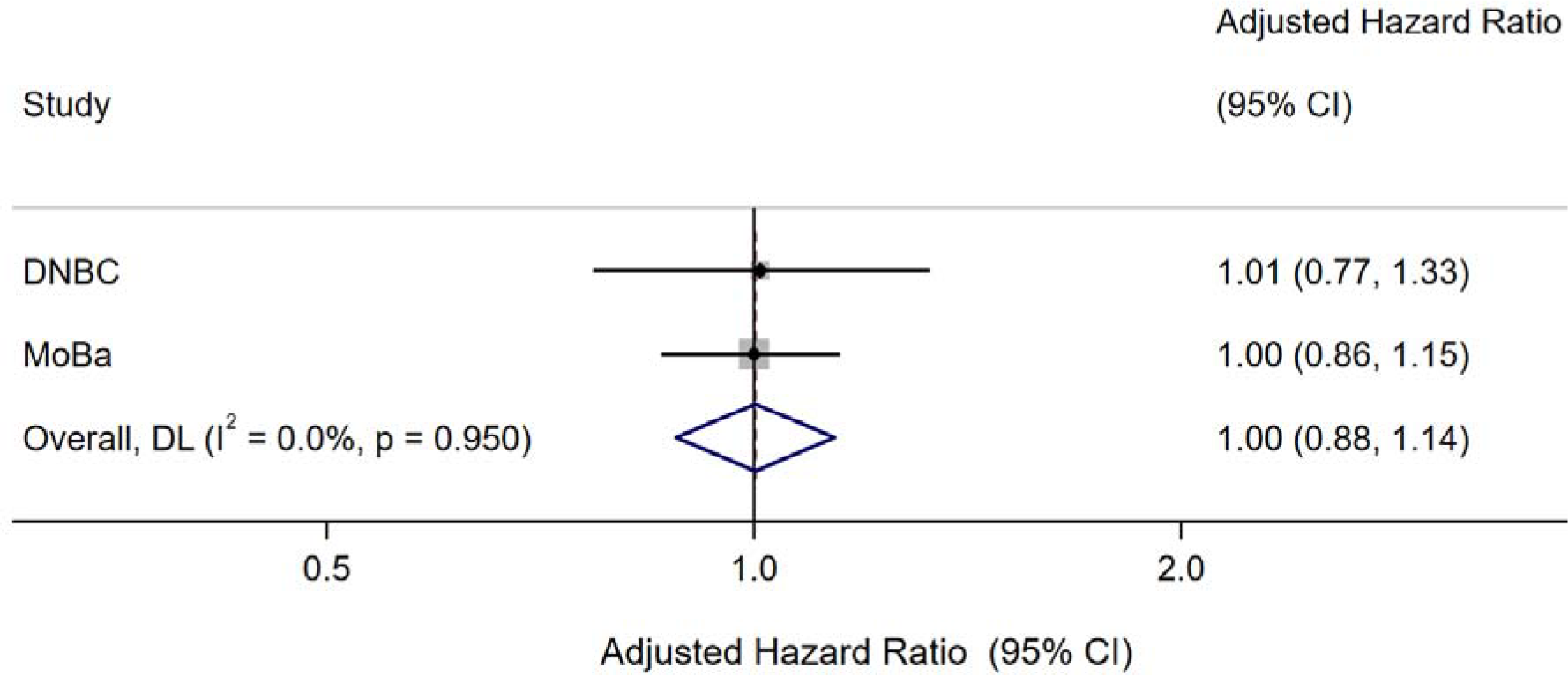
Maternal intake of EPA and DHA during pregnancy and risk of type 1 diabetes in children in DNBC and MoBa pregnancy cohorts. n=153,843 mother-child pairs, of whom n=634 children developed type 1 diabetes. Hazard ratios per gram per day intake of the sum of EPA and DHA from foods and supplements. Hazard ratios were adjusted for maternal parity, maternal pre-pregnancy BMI, maternal smoking during pregnancy, maternal type 1 diabetes, offspring sex, duration of breastfeeding and maternal education. Abbreviations: EPA: Eicosapentaenoic acid; DHA: Docosahexaenoic acid; DNBC: The Danish National Birth Cohort; MoBa (Norwegian acronym for): The Norwegian Mother, Father and Child Cohort Study; BMI: Body Mass Index; DL: DerSimonian and Laird random effects meta-analysis.

Robustness analyses also supported this conclusion, including adjustment for additional variables such as maternal vitamin D intake (Supplementary Table 3). Also, there were no deviation from linearity or any sign of a threshold effect from categorical analysis (adjusted hazard ratio comparing upper to lower fifth of intake was 1.19 (95% CI 0.81, 1.74) in DNBC and 0.84 (0.58, 1.23) in MoBa, Supplementary Table 3). Furthermore, secondary analyses showed no association of pregnancy intake of fish, or the common dietary 18-carbon chain omega-3 fatty acid alpha-linolenic acid, with risk of type 1 diabetes in the children (Supplementary Table 3).

## Discussion

Our data showed a complete lack of association between EPA and DHA intake during pregnancy and risk of type 1 diabetes, consistent in the two cohorts, with relatively narrow confidence intervals.

Our results are consistent with those in a high-genetic risk birth cohort from Finland (11). Our results therefore confirm and extend this lack of association in larger, general population cohorts. Another cohort with genetically susceptible children, although not investigating fatty acids directly, found no association between maternal fish intake and risk of type 1 diabetes in the children (14). Our cohorts were population based and assessed dietary intake including dietary supplements during pregnancy. We did not have biomarkers of EPA and DHA in our study, but the ability of the food frequency questionnaires to quantify n-3 fatty acids were in both cohorts validated against biomarkers (15, 16). A nested case-control study prospective study from Norway found no association with offspring type 1 diabetes of EPA and DHA in the phospholipid fraction of maternal serum collected in late pregnancy (17). While biomarkers are not influenced by recall and self-report, they are not without problems. Storage and handling of samples may lead to oxidation. Furthermore, biomarkers may be influenced by fasting or recent meals, as well as genetic and other factors regulating metabolism of fatty acids, depending on type of specimen and assay methods used (18). Common variants in the fatty acid desaturase (*FADS1*/*FADS2*/*FADS3*) gene cluster are associated with lower efficiency of conversion of the dietary precursor fatty acids alpha-linolenic acid to EPA and DHA and blood levels of these fatty acids (1). In the asthma prevention trial, both baseline blood levels of EPA and DHA and a genetic variant associated with lower blood levels of these fatty acids, were associated with a stronger relative effect of fish oil supplementation on prevention of asthma (10). On the other hand, studies of other disease outcomes in adults have not found consistent interactions between intake of EPA or DHA and genetic variants in the FADS gene cluster on cardiometabolic disease outcomes (1). Future studies of maternal n-3 fatty acid intake in pregnancy in relation to childhood type 1 diabetes could consider including genetic variants in the FADS gene cluster influencing conversion of alpha-linolenic acid to EPA and DHA.

We cannot exclude the possibility that intakes higher than that observed in our studies may show an association. Still, analysis of quintiles did not suggest any threshold effect. We adjusted for pregnancy intake of vitamin D in robustness analyses, even though previous evidence suggests no association with type 1 diabetes (19). Unmeasured confounding, for instance from toxicants in fatty fish may have influenced our results. However, there is currently no strong evidence supporting an association with such toxicants and type 1 diabetes (3) and we do not believe toxicants have confounded our results substantially. Given the largely Scandinavian origin or our participants, we believe our results are generalizable to other European origin populations, but not necessarily to populations with large proportions of people of other ancestries. Finally, our results do not exclude a potential effect of EPA and DHA intake in children rather than in their pregnant mothers.

In a setting where primary prevention trials are extremely expensive and time consuming (20), we believe our results together with the evidence discussed above have the clear implication that a trial of EPA and DHA during pregnancy to prevent type 1 diabetes in the offspring should not be prioritized. In conclusion, the hypothesis that a higher maternal omega-3 fatty acid intake during pregnancy reduce that risk of offspring type 1 diabetes was not supported.

## Supporting information

STROBE checklist

## Data Availability

Computer codes for the statistical analyses and aggregated data are available from the corresponding authors upon request to the corresponding author. The consent given by the participants in the two cohorts does not open for storage of individual level data in repositories or journals. Access to individual level data sets from the MoBa database requires an application, approval from The Regional Committee for Medical and Health Research Ethics in Norway, and an agreement with MoBa and the endpoint registries, which can be obtained by applying at https://helsedata.no/en/ (for more information, see the MoBa website https://www.fhi.no/en/ch/studies/moba/, or). Data from the Danish National Birth Cohort (DNBC) used in this study is managed by The DNBC Secretariat at Statens Serum Institut in Copenhagen, Denmark, and can be made available to researchers, provided approval from The DNBC organization, compliance with the EU General Data Protection Regulation (GDPR) and approval from the data owner (Statens Serum Institut). Researchers who wish to apply for access to data sets for replication purposes should apply through the DNBC Secretariat (see www.dnbc.dk). Access to data sets requires approval from the local Data Protection Agency (in a Danish context; "Fortegnelsen") at the researcher's institution and an agreement with the DNBC at Statens Serum Institut.

## Acknowledgements

We are grateful to all the participating families in Denmark and Norway who take part in these on-going cohort studies.

## Author contributions

SFO, LCS, NALB conceptualized the study. NALB and LCS drafted the protocol and a detailed analysis plan with input from SFO, AAB, MS, TIH, CG, GT, KS, and ALB. SFO, TIH, and CG were responsible for the dietary database in DNBC, and ALB in MoBa. JS (in DNBC), TS, PRN, and GJ (in MoBa) were responsible for the type 1 diabetes case validation and the diabetes registries. TIH and AAB carried out the statistical analyses of DNBC data, while NALB and LCS did the statistical analysis of the MoBa data and the meta-analyses. KS and GT contributed with data linkage and preparation including data cleaning and categorization in MoBa data. NALB and LCS drafted the manuscript and wrote the paper with input from all authors. SFO and LCS had the primary responsibility for the final content. All authors read and approved the final manuscript.

