## Supplementary for "Long chain omega-3 fatty acid intake in pregnancy and risk of type 1 diabetes in the offspring: Two large Scandinavian pregnancy cohorts – MoBa and DNBC"

Content

### Supplementary Methods

#### Population and Study Design

The Danish National Birth Cohort (DNBC) recruited pregnant women from 1996-2002 and the Norwegian Mother, Father and Child Cohort Study (MoBa) from 1999–2008. These are nation-wide prospective population-based pregnancy cohorts (1, 2). In Denmark, approximately 35% of all pregnant women were recruited into the DNBC corresponding to 101,042 pregnancies. In MoBa, approximately 40% of all pregnant women were recruited and in total, the cohort includes 95,200 mothers and 114,500 children. DNBC and MoBa together include 153,843 mother-child pairs with prospectively collected intake of EPA and DHA during pregnancy included in the current analyses (see Main Figure 1 Flow chart).

In the DNBC, women were recruited during their first antenatal visit to the general practitioner (gestational week ~6), whilst in MoBa, pregnant women received an invitation letter prior to the first ultrasound examination in gestational week ~18. Participants from the DNBC included in this study were women who answered a food frequency questionnaire (FFQ) in mid-pregnancy and participated in two telephone interviews on behavioral factors and health in gestational weeks 12 and 30. The MoBa participants who were included in this study were women who answered a questionnaire on behavioral factors and health in gestation week ~15 and an FFQ in week 22. The MoBa FFQ was introduced in 2002 and used throughout the recruitment period, while women participating in years 1999-2002 received a different FFQ that was not compatible with the questionnaire developed specifically for pregnant women.

#### Ethics

Written informed consent was obtained from all participants in both cohorts. The establishment of the cohort and data collection in MoBa were previously based on a license from the Norwegian Data Protection Agency and approval from The Regional Committee for Medical Research Ethics (S-95113/S-97045). They are now based on regulations related to the Norwegian Health Registry Act. The specific research question in the current sub-project was approved by the Regional Ethics Committee South-East C (2013/144/REK). The regional scientific ethics committee for the municipalities of Copenhagen and Frederiksberg approved all DNBC study protocols (27 August 2013: H-2-2013-108). All procedures were in accordance with the Declaration of Helsinki.

#### Inclusions and exclusions

In the DNBC, dietary information was available for a total of 70,183 pregnancies. Of those, 406 pregnancies were excluded due to unrealistically low (≤2.5 MJ) or high (>25.0 MJ) energy intake. Exclusion of further 1655 women with multiple pregnancies resulted in 68,122 pregnancies being included in our final analyses. The MoBa held information on 87,070 pregnancies with available maternal dietary information, and 1347 were excluded due to implausible energy intakes. The combined number of mother-child pairs available for analyses from both cohorts was 153,843 (main Figure 1 flow chart)

#### Dietary exposure variables

Dietary assessment methods in both cohorts have been described elsewhere (3). In both cohorts, the FFQs were designed and validated specifically for pregnant women (4-7), including validation of n-3 fatty acids against biomarkers in both cohorts (4, 6). Reported frequencies of food intake in the FFQs were scaled into amounts (g/day), by using assumptions of standard portion sizes, gathered from respective national dietary surveys, and by defining recipes for composite foods. The amount of energy and nutrients from each food item was then estimated based on linkage to the respective National Food Composition Table. The total amount of energy, intake of macronutrients and micronutrients was then estimated by aggregating the contribution from all food items. Specifically, long-chain n-3 polyunsaturated fatty acids (LCn3PUFAs) eicosapentaenoic acid (EPA) and docosahexaenoic acid (DHA), alpha-linolenic acid (ALA) and intake of total fish, lean and fatty fish were estimated in g/day. Portion sizes used for fish and fish products including cold cuts and spreads were similar in the two FFQs and the same software for nutrient calculations was used.

In addition, use of fish oil supplements was from the DNBC based on maternal report in structured telephone interviews conducted in gestational week 30 and six months postpartum. We also assessed the intake of all supplements, including fish oils, as reported when the women filled out the FFQ in gestational week 25. In MoBa, use of cod-liver oil/fish oil supplements was reported in the baseline questionnaire answered in gestational week 15 and covered three intervals prior to pregnancy (26-9 weeks, 8-5 weeks, and 4-0 weeks) and four intervals during pregnancy (0-4 weeks, 5-8 weeks, 9-12 weeks, 13+ weeks). In addition, use of supplements was assessed in detail in the FFQ in mid-pregnancy and converted to daily intakes of EPA and DHA based on information about brand, frequency, and amount (8, 9).

#### Outcome: type 1 diabetes

The clinical diagnosis of type 1 diabetes was defined as the first day of insulin treatment in accordance with the EURODIAB criteria. This information was obtained from the Norwegian and the Danish Childhood Diabetes Registries (10, 11) with additional linkage to the Norwegian Patient Registry (NPR) for MoBa participants to confirm the diagnosis. The most updated register linkage was included was end of 2019 for DNBC and April 15, 2018 for MoBa.

#### Covariates

Based on the a priori statistical analyses plan the following covariates were included in adjusted models: Maternal parity (12, 13), maternal pre-pregnancy body mass index (BMI) (14), maternal smoking during pregnancy (never/sometimes/daily) (15, 16), maternal type 1 diabetes (17, 18), offspring sex (17, 18), duration of breastfeeding (19, 20) and maternal education (21, 22) (categorized as in Supplementary Table 2).

The basis for the selection of covariates was to include in the primary analysis factors relatively robustly associated with risk of type 1 diabetes in previous studies, and at the same time were suspected to be associated with exposure (maternal intake of DHA+EPA) (23). We also considered availability of reasonably well measured variables and potential bias amplification if including strong predictors of exposure that were not predictors of offspring type 1 diabetes (23, 24). There were some exceptions from this principle: offspring sex which is associated with offspring type 1 diabetes, but not likely to influence exposure, and breastfeeding duration has not been associated with risk of type 1 diabetes in large prospective studies including MoBa and DNBC (25), but previous experience is that many readers prefer to see analyses adjusted for these variables. Similarly, socioeconomic status indicators such as maternal education is not robustly associated with type 1 diabetes, but is commonly adjusted for (21), and tended to be associated with type 1 diabetes in a large Norwegian cohort (22).

In robustness (sensitivity-) analyses presented in Supplementary Table 3 we additionally adjusted for the following variables: caesarean section (robustly but weakly associated with type 1 diabetes (18, 26, 27)), birth weight (robustly, but very weakly associated with type 1 diabetes, and likely to be a weak mediator rather than a confounder (28, 29)), maternal energy intake (not known to be associated with type 1 diabetes, but likely positively associated with exposure, and by some considered important to assess nutrient density, although we consider the absolute exposure most relevant in our setting, rather than nutrient density or substitution (31)), maternal intake of vitamin D from the diet and supplements combined (tend to be associated with exposure due to same food sources, and often stated to be associated with risk of type 1 diabetes despite the clear lack of association between pregnancy- or newborn vitamin D status in MoBa, DNBC (32) and several other large cohort studies (18, 33, 34)).

#### Statistical Analysis

In our primary analyses the association between maternal intake of LCn3PUFAs and offspring risk of type 1 diabetes was assessed using Cox proportional hazard models to estimate hazard ratios (HRs) with 95% confidence intervals (CIs) in each cohort separately. In absence of previous data to support any particular dose-response relationship, we hypothesized a log-linear association between exposure and outcome and used the exposure as a continuous variable. We adjusted for covariates listed above, and missing information on covariates were imputed using multiple imputation (m=5). Offspring age from birth up to end of 2019 was used in DNBC, and April 15^th^ 2018 for MoBa, as the underlying timescale censoring if death or emigration occurred. Among the MoBa children included in the analysis, the mean and maximum follow-up time was 12.3 years and 16.0 years respectively, and the total number of follow-up time was 1,051,628 person-years. Among the DNBC children included in the analysis, the mean and maximum follow-up time was 17.2 years and 20.4 years respectively, and the total number of follow-up time was 1,171,698 person-years. Because women could participate in the study repeatedly through different pregnancies, we used a robust sandwich covariance matrix estimate to account for interdependent observations. The proportional hazards assumption was evaluated by inspecting the Schoenfeld residuals. We used a random effect meta-analysis to pool the hazard ratios from the two cohorts (35) using the *metan* package in Stata version 17.

In additional robustness analyses (all presented in Supplementary Table 3) we assessed the linearity assumption in the primary analysis by also doing categorical analyses with the main exposure divided in fifths (quintiles), and we did complete case analyses only including subjects with no missing data on any covariates.

In secondary analyses (presented in Supplementary Table 3) the association of maternal intake of fish oil supplements, fatty fish, lean fish, and intake of alpha-linolenic acid were examined in relation to offspring risk of type 1 diabetes.

### Supplementary Tables

|  | **DNBC**  **(n=68 122)** | **MoBa**  **(****n=85 721)** |
| --- | --- | --- |
| **Sum of EPA and DHA from the diet (g/day)*** |  |  |
| Median (IQR) | 0.26 (0.15, 0.44) | 0.56 (0.32, 1.07) |
| Mean (SD) | 0.33 (0.29) | 0.81 (0.71) |
| **EPA+DHA from supplements (g/day)†** | (no. of users = 2469) | (no. of users = 58285) |
| Median (IQR) | 2.0 (0.5, 2.8) | 0.29 (0.14, 0.80) |
| Mean (SD) | 2.0 (1.7) | 0.55 (0.60) |
| **Fatty fish (g/day)** |  |  |
| Median (IQR) | 0.0 (0.0, 5.3) | 8.3 (3.8, 15.6) |
| Mean (SD) | 3.5 (5.7) | 12.2 (14.0) |
| **Lean fish (g/day)** |  |  |
| Median (IQR) | 6.4 (2.5, 11.9) | 18.7 (10.8; 27.9) |
| Mean (SD) | 8.3 (8.3) | 20.4 (13.5) |
| **Alpha-linolenic acid (g/d)** |  |  |
| Median (IQR) | 1.9 (2.5) | 1.5 (1.1, 1.9) |
| Mean (SD) | 2.1 (0.9) | 1.6 (0.7) |

#### **Supplementary Table 1.** Estimated intake of sum EPA and DHA, fish and alpha-linolenic acid among pregnant women in the DNBC and the MoBa cohorts

* Maternal intakes of EPA and DHA were strongly correlated (Spearman’s correlation coefficient=0.98, p <0.001).

† Among users of supplements.

Abbreviations: IQR: Interquartile range (25^th^ percentile, 75^th^ percentile); SD: Standard Deviation; DNBC: Danish National Birth Cohort; MoBa (Norwegian acronym for): Norwegian Mother, Father and Child Cohort Study; EPA: eicosapentaenoic acid; DHA: Docosahexaenoic acid.

|  | **DNBC**  **(n=68 122)** | **MoBa**  **(n=85 721)** |
| --- | --- | --- |
| **Maternal Characteristics** |  |  |
| *Parity* [n, (%)] |  |  |
| 0 | 31940 (48%) | 39158 (46%) |
| 1 | 23987 (36%) | 30345 (35%) |
| 2 or more | 10073 (16%) | 16218 (19%) |
| Missing [n] | 2122 | 0 |
| *Education* [n, (%)] |  |  |
| Less than 9th grade (DNBC)/Less than high school (MoBa) | 3426 (7%) | 6089 (7%) |
| 10th grade (DNBC)/High school (MoBa) | 13212 (26%) | 24523 (29%) |
| Technical school (DNBC)/Up to 4 years of college (MoBa) | 7022 (14%) | 35043 (41%) |
| High School or more (DNBC)/More than 4 years of college (MoBa) | 27478 (54%) | 19677 (23%) |
| Missing [n] | 16984 | 389 |
| *Smoking during pregnancy (daily or sometimes)* [n, (%)] |  |  |
| No | 50987 (76%) | 77946 (91%) |
| Yes | 16440 (14%) | 7314 (8.6%) |
| Missing [n] | 695 | 461 |
| *Maternal type 1 diabetes* |  |  |
| No | 67927 (99.7%) | 85379 (99.6%) |
| Yes | 195 (0.3%) | 342 (0.4%) |
| *Pre-pregnancy BMI* [n, (%)] |  |  |
| <20 | 11002 (18%) | 10351 (12%) |
| 20- <25 | 35314 (59%) | 46705 (56%) |
| 25 to <30 | 12517 (21%) | 18420 (22%) |
| $\geq30$ | 1393 (2%) | 8074 (10%) |
| Missing [n] | 7896 | 2171 |
| *Breastfeeding duration in months* [n, (%)] |  |  |
| <4 | 10309 (21%) | 10330 (14%) |
| 4 - <6 | 9031 (18%) | 6594 (9%) |
| ≥6 | 29814 (61%) | 59218 (78%) |
| Missing [n] | 18969 | 9579 |
| **Offspring characteristics** |  |  |
| *Sex* [n, (%)] |  |  |
| Boys | 34742 (51%) | 43907 (51%) |
| girls | 33380 (49%) | 41814 (49%) |

#### **Supplementary Table 2.** Characteristics of study participants in the DNBC and the MoBa cohort.

**Supplementary Table 3**. Robustness and secondary analyses in the DNBC (n=68,122) and the MoBa cohorts (n=85,721)^1^

|  | **Unadjusted DNBC** | **Adjusted DNBC^2^** | **Unadjusted MoBa** | | **Adjusted MoBa^2^** | **Adjusted, pooled^3^** |
| --- | --- | --- | --- | --- | --- | --- |
|  | HR (95% CI) | HR (95% CI) | HR (95% CI) | | HR (95% CI) | HR (95% CI) |
| **Primary analyses*** | |  |  | |  |  |
| Sum of EPA and DHA (g/d) | 0.98 (0.73, 1.31) | 1.01 (0.77, 1.33) | 0.98 (0.85, 1.14) | | 1.00 (0.86, 1.15) | 1.00 (0.88, 1.14) |
| **Categorical analysis to assess potential non-linearity**^4^  (Quintiles of sum of EPA and DHA intake) | | |  | |  |  |
| 1^st^ quintile (reference) | 1.00 (reference) | 1.00 (reference) | 1.00 (reference) | | 1.00 (reference) |  |
| 2^nd^ quintile | 1.13 (0.79, 1.61) | 1.36 (0.94, 1.95) | 0.92 (0.66, 1.27) | | 0.83 (0.57, 1.20) |  |
| 3^rd^ quintile | 0.96 (0.66, 1.40) | 1.10 (0.74, 1.61) | 0.98 (0.71, 1.35) | | 0.84 (0.58, 1.23) |  |
| 4^th^ quintile | 0.88 (0.60, 1.29) | 1.01 (0.67, 1.50) | 1.04 (0.75, 1.42) | | 0.91 (0.62, 1.33) |  |
| 5^th^ quintile | 1.02 (0.71, 1.48) | 1.19 (0.81, 1.74) | 0.89 (0.64, 1.24) | | 0.84 (0.58, 1.23) |  |
| ***Robustness analyses****:* | | |  | |  |  |
| Complete case analysis:  (DNBC n=42221, MoBa n= 73707)^2^ | | 0.63 (0.41, 0.98) |  | | 0.96 (0.82, 1.13) | 0.82 (0.55, 1.22) |
| Excluding multiple pregnancies in MoBa (n=3119), complete case^2^ | | |  | | 0.97 (0.83, 1.14) |  |
| *Additional adjustments of primary analysis for* | |  |  | |  |  |
| -Energy intake |  | 0.92 (0.65, 1.30) |  | | 0.95 (0.80, 1.12) | 0.94 (0.81, 1.10) |
| -Vitamin D from diet and supplements |  | 0.94 (0.70, 1.27) |  | | 0.95 (0.79, 1.14) | 0.95 (0.81, 1.11) |
| -Mode of delivery (vaginal vs Cesarean) |  | 1.01 (0.77, 1.33) |  | | 0.95 (0.81, 1.12) | 0.97 (0.84, 1.11) |
| -Birth weight |  | 1.01 (0.76, 1.33) |  | | 0.96 (0.81, 1.12) | 0.96 (0.84, 1.11) |
| **Secondary analyses**^1^ |  |  |  | |  |  |
| Fish oils Suppl (Yes vs No) | 1.32 (0.76, 2.30) | 1.34 (0.78, 2.36) | 0.84 (0.68, 1.04) | | 0.79 (0.62, 1.01) | 0.97 (0.58, 1.60) |
| Fatty fish, continuous (per 10 g/d) | 0.99 (0.80, 1.22) | 1.02 (0.84, 1.25) | 1.00 (0.93, 1.08) | | 0.99 (0.91, 1.08) | 1.00 (0.92, 1.08) |
| Lean fish, continuous (per 10 g/d) | 1.05 (0.91, 1.20) | 1.07 (0.94, 1.22) | 0.98 (0.91, 1.06) | | 0.96 (0.87, 1.05) | 1.01 (0.89, 1.16) |
| Alpha-linolenic acid (per g/d) | 1.03 (0.91, 1.17) | 1.04 (0.92, 1.19) | 1.07 (0.92, 1.24) | | 1.04 (0.89, 1.21) | 1.04 (0.94, 1.15) |

^*^ Same as shown in main Figure 2, for comparison with robustness analyses (sum of EPA and DHA intake during pregnancy, continuous in units g/day), with missing data for covariates imputed as described in Supplementary Methods.

^1^ DNBC (n=68,122, of whom 279 developed type 1 diabetes) and the MoBa cohort1 (n=85,721 of whom 355 developed type 1 diabetes). ^2^ Adjusted for maternal pre-pregnancy type 1 diabetes, maternal age, maternal education, maternal body mass index, parity, smoking during pregnancy and duration of breastfeeding without imputation of missing data for covariates. ^3^ Q-tests for heterogeneity between DNBC and MoBa suggested no significant heterogeneity: Extra adjustment for total energy intake – P(het.)=0.87; Extra adjustment for vitamin D intake – P(Het)=0.95; extra adjustment for mode of delivery - P(het.)=0.71; Extra adjustment for birth weight - P(het.)=0.71; Complete case analysis – P(het.)=0.075; Fish oil supplements (yes/no) - P(het.)=0.087; fatty fish intake - P(het.)=0.76; lean fish intake - P(het.)=0.18; Alpha-linolenic acid – P(het.)=0.99. ^4^ These results were not pooled because the absolute cut-off values in the two studies differed. The purpose to assess possible deviations from linearity including possible threshold associations is done best by assessing each cohort separately. Quintile cut-offs (and mean) for the sum of EPA and DHA intake (g/d) in MoBa were 1: <0.28 (mean 0.18), 2: 0.28-0.45 (mean 0.36), 3: 0.45-0.72 (mean 0.57), 4: 0.72-1.2 (mean 0.95), 5: ≥1.2 (mean 1.98).

Quintile cut-offs and mean in DNBC were 1: <0.18 (mean 0.12), 2: 0.18-0.30 (mean 0.24), 3: 0.30-0.43 (mean 0.36), 4: 0.43-0.64 (mean 0.52), 5: ≥ 0.64 (mean 0.96)

Abbreviations: DNBC: Danish National Birth Cohort; MoBa (Norwegian acronym for): Norwegian Mother, Father and Child Cohort Study; EPA: eicosapentaenoic acid; DHA: Docosahexaenoic acid; HR: Hazard Ratio; CI: Confidence Interval.
