## Supplementary material for "Long chain omega-3 fatty acid intake in pregnancy and risk of type 1 diabetes in the offspring: Two large Scandinavian pregnancy cohorts – MoBa and DNBC": STROBE checklist

STROBE Statement—Checklist of items that should be included in reports of ***cohort studies***

|  | Item No | Recommendation |
| --- | --- | --- |
| **Title and abstract** | 1 | (*a*) Indicate the study’s design with a commonly used term in the title or the abstract: p1 |
|  |  | (*b*) Provide in the abstract an informative and balanced summary of what was done and what was found p2 |
| Introduction | | |
| Background/rationale | 2 | Explain the scientific background and rationale for the investigation being reported p3 |
| Objectives | 3 | State specific objectives, including any prespecified hypotheses p3 |
| Methods | | |
| Study design | 4 | Present key elements of study design early in the paper p4 |
| Setting | 5 | Describe the setting, locations, and relevant dates, including periods of recruitment, exposure, follow-up, and data collection p4+Suppl material Methods p1-3 |
| Participants | 6 | (*a*) Give the eligibility criteria, and the sources and methods of selection of participants. Describe methods of follow-up p4+Suppl material Methods p1-3 |
|  |  | (*b*) For matched studies, give matching criteria and number of exposed and unexposed n.a. |
| Variables | 7 | Clearly define all outcomes, exposures, predictors, potential confounders, and effect modifiers. Give diagnostic criteria, if applicable p4+Suppl material Methods p1-4 |
| Data sources/ measurement | 8* | For each variable of interest, give sources of data and details of methods of assessment (measurement). Describe comparability of assessment methods if there is more than one group p4+Suppl material Methods p1-4 |
| Bias | 9 | Describe any efforts to address potential sources of bias p4+Suppl material Methods p1-4 |
| Study size | 10 | Explain how the study size was arrived at p4+Suppl material Methods p1-4+Fig 1 |
| Quantitative variables | 11 | Explain how quantitative variables were handled in the analyses. If applicable, describe which groupings were chosen and why p4+Suppl material Methods p1-4 |
| Statistical methods | 12 | (*a*) Describe all statistical methods, including those used to control for confounding –``– |
|  |  | (*b*) Describe any methods used to examine subgroups and interactions Suppl Table 3 (p7) |
|  |  | (*c*) Explain how missing data were addressed Suppl Mat. Methods p4 (multiple imputation for covariates), |
|  |  | (*d*) If applicable, explain how loss to follow-up was addressed (n.a. outomes linked to complete registries) |
|  |  | (*e*) Describe any sensitivity analyses Suppl material Methods p3-4 |
| Results | | |
| Participants | 13* | (a) Report numbers of individuals at each stage of study—eg numbers potentially eligible, examined for eligibility, confirmed eligible, included in the study, completing follow-up, and analysed Suppl material Methods p1-4 + Flow chart main Fig 1 |
|  |  | (b) Give reasons for non-participation at each stage Suppl material Methods p1-4 + Flow chart main Fig 1 |
|  |  | (c) Consider use of a flow diagram Main Fig 1 |
| Descriptive data | 14* | (a) Give characteristics of study participants (eg demographic, clinical, social) and information on exposures and potential confounders p5 + Suppl Tables 1-2 |
|  |  | (b) Indicate number of participants with missing data for each variable of interest Suppl Table 2 |
|  |  | (c) Summarise follow-up time (eg, average and total amount) Suppl Mat. Methos p. 4 |
| Outcome data | 15* | Report numbers of outcome events or summary measures over time p5 + Fig 1 |
| Main results | 16 | (*a*) Give unadjusted estimates and, if applicable, confounder-adjusted estimates and their precision (eg, 95% confidence interval). Make clear which confounders were adjusted for and why they were included Main Fig 1, Suppl Table 3 + Suppl Methods p3-4 |
|  |  | (*b*) Report category boundaries when continuous variables were categorized Footnote of Suppl Table 3 |
|  |  | (*c*) If relevant, consider translating estimates of relative risk into absolute risk for a meaningful time period : n.a. (because RR=1.00, null/no difference |
| Other analyses | 17 | Report other analyses done—eg analyses of subgroups and interactions, and sensitivity analyses p5 and Suppl Table 3 |
| Discussion | | |
| Key results | 18 | Summarise key results with reference to study objectives p6 |
| Limitations | 19 | Discuss limitations of the study, taking into account sources of potential bias or imprecision. Discuss both direction and magnitude of any potential bias p6-7 |
| Interpretation | 20 | Give a cautious overall interpretation of results considering objectives, limitations, multiplicity of analyses, results from similar studies, and other relevant evidence p7 |
| Generalisability | 21 | Discuss the generalisability (external validity) of the study results p7 |
| Other information | | |
| Funding | 22 | Give the source of funding and the role of the funders for the present study and, if applicable, for the original study on which the present article is based p9 |

*Give information separately for exposed and unexposed groups.

**Note:** An Explanation and Elaboration article discusses each checklist item and gives methodological background and published examples of transparent reporting. The STROBE checklist is best used in conjunction with this article (freely available on the Web sites of PLoS Medicine at http://www.plosmedicine.org/, Annals of Internal Medicine at http://www.annals.org/, and Epidemiology at http://www.epidem.com/). Information on the STROBE Initiative is available at http://www.strobe-statement.org.
